# Safety and feasibility of research lumbar puncture in Huntington’s disease: the HDClarity cohort and bioresource

**DOI:** 10.1101/2021.07.30.21261340

**Authors:** Filipe B Rodrigues, Gail Owen, Swati Sathe, Elena Pak, Dipinder Kaur, Anka G Ehrhardt, Sherry Lifer, Jenny Townhill, Katarzyna Schubert, Blair R Leavitt, Mark Guttman, Jee Bang, Jan Lewerenz, Jamie Levey, for the HDClarity Investigators, Cristina Sampaio, Edward J Wild

## Abstract

**Background:** Biomarkers are needed to monitor disease progression, target engagement and efficacy in Huntington’s disease (HD). Cerebrospinal fluid (CSF) is an ideal medium to research such biomarkers due to its proximity to the brain.

**Objectives:** To investigate the safety and feasibility of research lumbar punctures (LP) in HD.

**Methods:** HDClarity (NCT02855476) is an ongoing international biofluid collection initiative built on the Enroll-HD platform, where clinical assessments are recorded. It aims to recruit 1,200 participants. Biosamples are collected following an overnight fast: blood via venipuncture and CSF via LP. Participants are healthy controls and HD gene expansion carriers across the disease spectrum. We report on monitored data from February 2016 to September 2019.

**Results:** Of 448 participants screened, 398 underwent at least 1 sampling visit, of which 98.24% were successful (i.e. CSF was collected), amounting to 10,610mL of CSF and 8,200mL of plasma. In the total 572 sampling visits, adverse events were reported in 24.13%, and headaches of any kind and post-LP headaches in 14.86% and 12.24%, respectively. Frequencies were less in manifest HD; gender, age, body mass index and disease burden score were not associated with the occurrence of the events in gene expansion carriers. Headaches and back pain were the most frequent adverse events.

**Conclusions:** HDClarity is the largest CSF collection initiative to support scientific research into HD and is now established as a leading resource for HD research. Our data confirm that research LP in HD are feasible and acceptable to the community, and have a manageable safety profile.

## INTRODUCTION

Huntington’s disease (HD) is a progressive autosomal dominant genetic disease, which typically manifests in adulthood with movement disorder, cognitive decline, and psychiatric changes(1). Its overall survival after clinical diagnosis is around 20 years(2). There are currently no disease-modifying interventions available (3), but several clinical trials are underway and planned in the next few years to explore novel therapeutic approaches to treating this disease(4-6). In preparation for such trials, biomarkers are needed – especially prognostic, pharmacodynamic, efficacy and safety biomarkers.

Cerebrospinal fluid (CSF) is a favourable biofluid compartment for assessing HD biomarkers, owing to its proximity to the brain and consequent enrichment of CNS-derived products. Blood is less CNS-enriched but more accessible, and may provide relevant hints to monitor disease progression and assess response to treatments(7, 8), whilst being useful to help interpret findings in CSF. The CSF is also the compartment into which the first targeted huntingtin-lowering experimental therapeutic was delivered and the fluid in which its successful target engagement was assessed(9, 10).

There have been numerous reports of potential biofluid biomarkers in HD(11-13); however, many were assessed in small-scale cross-sectional studies and remain unvalidated. Even findings from larger studies with longitudinal designs(7, 8, 14) need to be replicated in a well-powered and standardized new sample set. New samples are also invaluable for the discovery of novel biomarkers and validation of modern analytical methods.

While CSF is generally sampled through a minimally invasive procedure, the safety and feasibility profile of lumbar punctures (LP), has not been systematically investigated in HD. In the general population, this procedure has a low risk of serious adverse events, such as CNS infection and bleeding. Post-LP back pain and headache are common but transient, and either have spontaneous resolution or need simple measures and reassurance. LP has been widely studied for research and clinical purposes in other neurodegenerative conditions, notably Alzheimer’s disease (AD), where older age and prominent generalised brain atrophy reduce the risk of the most common low-pressure syndromes(15-19). The HD population has important characteristics that could modify LP safety and feasibility profiles: relatively young age distribution compared to other dementias; degree of brain atrophy intermediate between healthy controls and AD patients; involuntary movements that could make LP more challenging; and a dysexecutive syndrome that could make recruitment, consent and toleration of procedure and adverse effects more difficult.

HDClarity (NCT02855476) commenced as a prospective nested CSF and blood collection initiative within Enroll-HD (https://enroll-hd.org)(20) with HD gene expansion carriers (HDGECs) and healthy control participants recruited from the main cohort to facilitate biomarker development in HD. Here we report the characteristics and experiences of the first 448 participants screened between February 2016 and September 2019. In addition, we examine the safety and feasibility of the study procedures in HDGEC and healthy controls, factors influencing complication risk, and quality control indicators of the collected CSF and plasma.

## MATERIALS AND METHODS

### Study protocol

The open-access HDClarity study protocol is available at http://hdclarity.net/study-information/(21). HDClarity (NCT02855476) was designed as a cross-sectional study with optional short-term resampling visits. It has since been extended to an annual collection for willing and eligible Enroll-HD participants at HDClarity sites to generate longitudinal samples and data.

### Study aims

The primary objective of HDClarity is to generate a high-quality CSF collection for evaluation of biomarkers and pathways that will enable the development of novel treatments for HD. The secondary objectives are to generate a high-quality plasma sample collection matching the CSF collections, which will also be used to evaluate biomarkers and pathways of relevance to HD research and development.

### Study design

HDClarity is a global longitudinal observational study, with the aim of enrolling 1,200 participants. All willing and eligible Enroll-HD participants at HDClarity sites are invited to participate in the HDClarity study.

The Enroll-HD study is a prospective longitudinal observational study that collects natural history data in HDGECs and healthy controls with core required assessments focused on neuropsychiatric, cognitive, motor and functional status conducted via a battery of validated and widely accepted measures. The Enroll-HD database includes clinical information on 24,391 participants (as of 21 Oct 2020) of which 17,734 are HDGECs, 2,406 are genotype unknown (but at risk of having inherited the expanded HD allele) and 4,251 are healthy controls. The mean age of HDGECs and healthy controls at enrolment into Enroll-HD was 49.1 years (range 11 – 92) and 48.8 years (range 18 to 91), respectively, with male to female ratio 1:1.16 and 1:1.51 respectively. Enroll-HD is conducted at 157 active sites in 19 countries across four continents.

Clinical and phenotypic data must be collected in the annual Enroll-HD visit within 2 months prior to the screening for HDClarity; otherwise the core assessments are repeated during HDClarity screening visit. Core Enroll-HD assessments include Unified Huntington’s Disease Rating Scale (UHDRS) Total Motor Score (TMS), Diagnostic Confidence Score (DCS), Total Functional Capacity (TFC), Independence Scale (IS), Problem Behaviours Assessment - short (PBA-S), Symbol Digit Modalities Test (SDMT), Stroop Word Reading (SWR), Stroop Color Naming (SCN), and Verbal Fluency Categorical (VFC)(22-26).

Participants who meet the HDClarity eligibility requirements at the screening visit, which includes testing platelet count and clotting function, return for a sampling visit within a month. Biosamples are collected between 8:00 and 10.30 am following an overnight fast: blood is obtained via venepuncture and CSF via LP. UHDRS TMS is repeated on the day of sampling. Participants are discharged following a period of observation and contacted within 72 hours to assess adverse events.

Approximately 20% of the HDClarity participants are invited to return for an optional short-term resampling visit approximately 4-8 weeks later. All participants are invited to return for annual visits.

### Ethical considerations

HDClarity is performed in accordance with the principles of the Declaration of Helsinki, and the International Conference on Harmonization and the World Health Organization Good Clinical Practice standards. All participating sites sought appropriate ethical approval in accordance with each specific country legislation (more details including Ethics Committees / Institutional Review Boards decisions in Supplementary Appendix 1). All reported participants gave informed consent prior to undertaking study procedures.

### Study population

Six participant groups are being recruited of which five are sub-categories of HDGEC (early pre-manifest, late pre-manifest, early manifest, moderate manifest and advanced manifest HD patients) in addition to healthy controls. Healthy controls have either no known family history of HD, or have had a negative genetic test for the HD CAG expansion (i.e. CAG < 36). All HDGEC participants need to have been tested locally for the huntingtin gene glutamine codon (CAG) expansion and have a CAG ≥40 if premanifest or ≥ 36 if manifest. Further details of subgroup characteristics are provided in **Error! Reference source not found**.. Enrolment is anticipated to be similar in each group.

Eligible participants are aged between 21 and 75 years, inclusive, and of both genders. They need to be able to provide informed consent or have a legal representative authorized to give consent on their behalf. Compliance with study procedures, including fasting, blood sampling and LP is required, as is participation in the Enroll-HD study(20).

The main exclusion criteria are: participation in a clinical drug trial within 30 days prior to the sampling visit or use of an investigational drug; significant medical, neurological or psychiatric co-morbidity likely to impair participant’s ability to complete study procedures, or likely to reduce the utility of the samples and data for studies of HD; clinical or laboratory bleeding and inflammatory abnormalities. For a detailed description please refer to the study protocol.

### Biosample collection, processing, and storage

To minimize inter-site and inter-sample variability, biosamples (i.e. CSF and blood) are collected and processed according to a standardized and pre-piloted protocol (see Supplementary appendix 2).

Briefly, local sites are provided with centrally sourced kits for CSF and blood collection and processing. A LP is performed using a 22G Whitacre atraumatic BD spinal needle. Up to 20 mL of CSF are collected into a 50 mL pre-cooled polypropylene collection tube on wet ice. Biosamples are transported to the laboratory on wet ice, with the exception of serum which is kept at room temperature, and sample processing starts within 15 minutes of collection.

Red and white cell counts are carried out in up to 200 µL of CSF onsite for safety and quality control purposes, and the remaining CSF is prepared and aliquoted into 300 µL cryovials. Plasma is prepared and aliquoted into 300 µL cryovials and serum into 1,500 µL cryovials. All biosamples are frozen at - 80°C and then shipped to a central biorepository using centrally sourced boxes with dry ice and temperature probes.

### Site recruitment

Larger Enroll-HD sites were prioritised for HDClarity study to facilitate recruitment. Subsequent sites were added after assessment for suitability based on experience, expertise and facilities, local ethical approval, legal approval, translation of materials and site training. The number of sites opened annually from 2016 to 2019 were 2, 7, 5 and 1, for a total of 15 sites.

### Data managing and statistical analysis

This analysis reports on fully monitored data from participants recruited from February 2016 to September 2019. All data were recorded on the Enroll-HD electronic data capture system. Data were remotely monitored by HDClarity central coordination team, and on-site by trained Enroll-HD data monitors. The final analysis dataset was queried for implausibilities, which were removed from the dataset and assumed as missing. No imputation procedures were used.

To describe study visits, the unit of analysis was the study visit irrespective of the fact that a proportion of participants who had annual sampling visits had more than 1 screening (n= 71; 15.85%) and/or sampling visits (n= 68; 17.09%). Participants with at least one successful sampling visit (i.e. where dura was pierced and CSF was collected, irrespective of amount of CSF) are characterized using data from the initial sampling visit. Adverse event data are analyzed for successful and unsuccessful (i.e. where a LP was attempted, but no CSF was collected) sampling visits. Headaches were defined as any kind of head pain, and post-lumbar puncture headaches (PLPH) as a headache secondary to a low CSF pressure syndrome as judged by the local site investigator.

Continuous variables were reported as mean ± standard deviations (SD); counts as median ± interquartile range (IQR), minimum and maximum; and categorical variables as absolute (n) and relative frequencies (%). Intergroup differences in continuous variables were tested with linear regression, and intergroup independence across categorical variable was examined with logistic regression. 2-sided Fisher exact tests were used for categories with zero events. We reported unadjusted p-values for the omnibus group membership main effect test and relevant contrasts.

To identify possible factors associated with LP success and post-LP complications, we examined the associations between the event of interest (i.e. LP success, adverse events, headache, PLPH) and exposures of interest using univariable mixed effects logistic regression, with the event as the dependent variable and participant as a random intercept. A multivariable model including study group, age, gender, body mass index (BMI; kg·m^-2^), and Disease Burden Score (DBS; [CAG-35.5]·age)(27) as fixed independent variables was used to report the adverse event frequency within each study group. We reported unadjusted p-values for the omnibus group membership main effect test and relevant contrasts. To study the impact of age, gender, BMI and DBS in HDGEC only these four variables were included in the model as fixed variables. DBS is a measure of cumulative exposure to HD pathology as a function of CAG repeat length and time exposed to the effects of the expansion. The output was reported as odds ratio (OR) and 95% confidence interval (95% CI). P-values were not adjusted for multiple comparisons, and data analysis was performed with the statistical package StataMP 16 (StataCorp, Texas, USA).

## RESULTS

### Participant and visit characteristics

In the first 43 months of the study, 448 participants were screened, of whom 398 went on to have one or more sampling visits. 459 successful sampling visits were completed at 15 study sites, followed by 101 successful short-term repeat sampling visits. Overall, 560/572 (97.9%) sampling visits were deemed successful (Figure 1 and **Error! Reference source not found**.). Characteristics of those who underwent successful sampling are shown in

**Figure 1.**
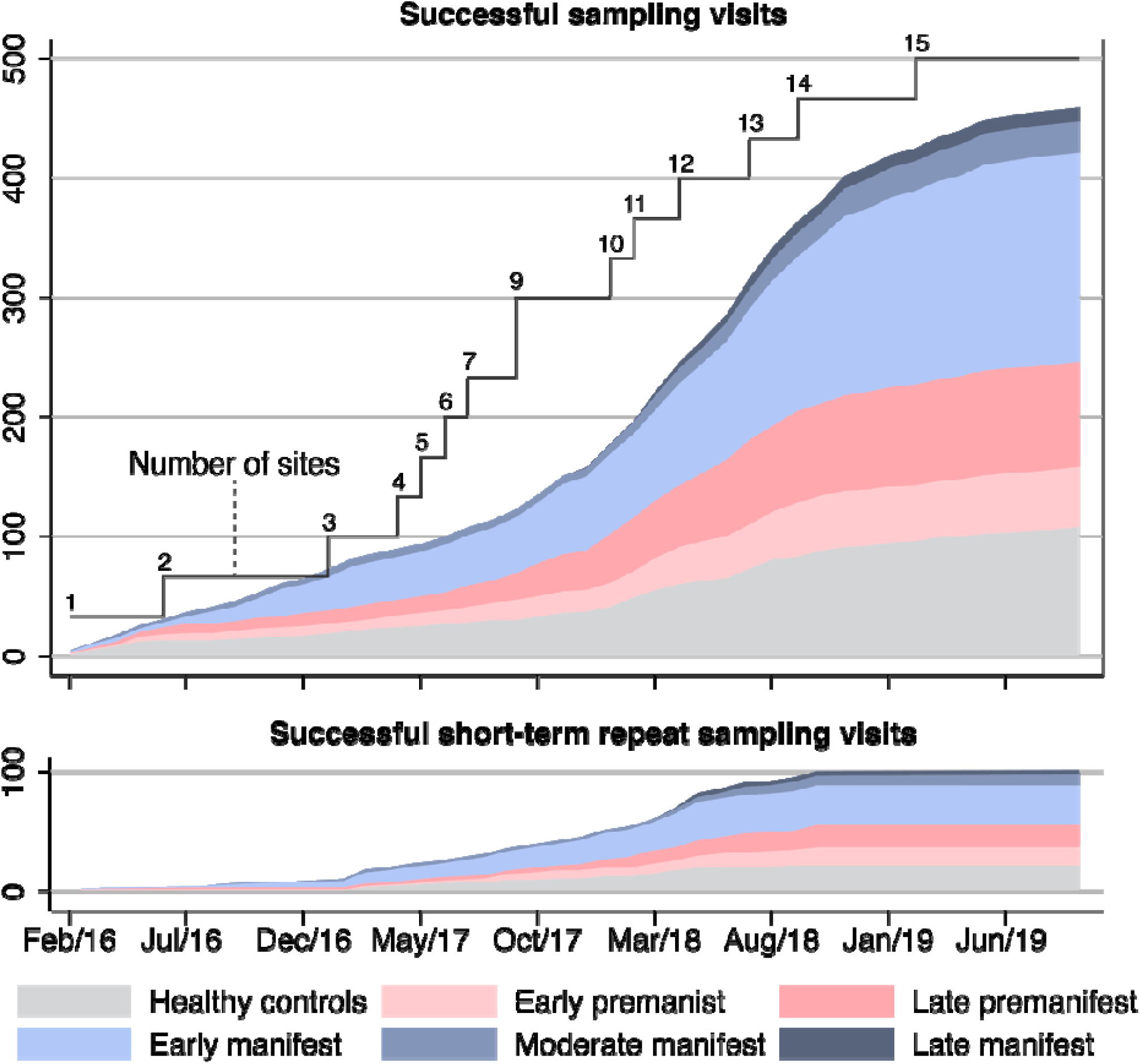
Successful sampling and short-term repeat sampling visits over time. Successful visits were defined as when dura was pierced and CSF was collected, irrespective of amount of CSF. Short-term repeat sampling visits were paused across most participant groups in early 2019 when the initial target numbers were reached; hence no fully monitored visits of this kind were captured in the dataset from January to September 2019.

Table 1 and a comparison between participants’ characteristics at successful and unsuccessful visits in **Error! Reference source not found**..

*Table 1 –* ***Characteristics at first screening visit of participants who underwent at least 1 successful sampling visit***. *Successful visits were defined as when dura was pierced and CSF was collected, irrespective of amount of CSF. Continuous variables are reported as mean ± standard deviations. Categorical variables are reported as absolute and relative frequencies. BMI, body mass index; CAG, CAG repeat count; DBS, Disease Burden Score; UHDRS, Unified Huntington’s Disease Rating Scale; TMS, UHDRS Total Motor Score; TFC, UHDRS Total Functional Capacity; IS, UHDRS Independence Score; FA, UHDRS Functional Assessment; SWR, Stroop Word Reading test; SCN, Stroop Color Naming test; SDMT, Symbol Digits Modality Test; VFC, Verbal Fluency Categorical; n/a, not applicable*.

### Adverse events

In 572 sampling and repeat sampling visits, one or more adverse events were reported in 138 visits (24.13%, Table 2, **Error! Reference source not found**.); in 86 (62.32%) visits they were mild, in 51 (36.96%) visits moderate and in 1 (0.72%) visit was severe. The median duration was 4 days (IQR 5, max 28, min 1). Overall, there were 189 reported adverse events (Table S4): 118 (62.43%) were mild, 70 (37.04%) were moderate and 1 (0.53%) was severe; and 152 (80.42%) were deemed “probably” related to the study procedure, 25 (13.23%) were “possibly” related, and 12 (6.35%) were unrelated. The most frequent side effect was headache, followed by back pain (**Error! Reference source not found**.). Headaches of any kind were reported after 85 visits (14.86%, Table 2). Of these, 50 (58.82%) were mild, 34 (40.00%) were moderate and 1 (1.18%) was severe. The median duration was 4 days (IQR 4, max 18, min 1). PLPH, defined by the local site investigator as headache secondary to a low CSF pressure syndrome, were reported after 70 visits (12.24%, Table 2, Table S3); 36 (51.43%) were mild, 33 (47.14%) were moderate and 1 (1.43%) was severe. The median duration was 5 days (IQR 4, max 18, min 1).

There was a single (0.17%) serious adverse event, where one female participant had to be admitted for a blood patch due to a prolonged, moderate intensity PLPH. A second blood patch was recorded following a prolonged non-serious PLPH of moderate intensity in female participant. In total there were 2 (0.35%) blood patches, both effective in relief of PLPH.

Participants with manifest HD had numerically fewer adverse events, headaches and PLPH than healthy controls; premanifest HD participants were not different from healthy controls, but given the low numbers of adverse events overall, these findings should be interpreted with caution (Table 2).

*Table 2 –* ***Frequency of adverse events per visit***. *Categorical variables are reported as absolute and relative frequencies. PLPH, post-lumbar puncture headache; HD, Huntington’s disease*.

We assessed whether age, gender, BMI and disease burden score – a measure combining age and CAG repeat length that is associated with clinical severity and degree of brain atrophy in HD(28, 29) – were predictors of PLPH. None of these variables were consistently associated with the occurrence of adverse events in HD mutation carriers in our dataset (Figure 2). The only predictor whose odds ratio deviated from 1.0 was gender: there was a somewhat higher odds of adverse events, headaches and PLPH in female gene expansion carriers (OR 1.57 for adverse events, OR 1.34 for headache and 1.12 for PLPH), but the 95% confidence intervals included 1.0 (0.64-3.89, 0.53-3.39 and 0.38-3.33 respectively).

**Figure 2.**
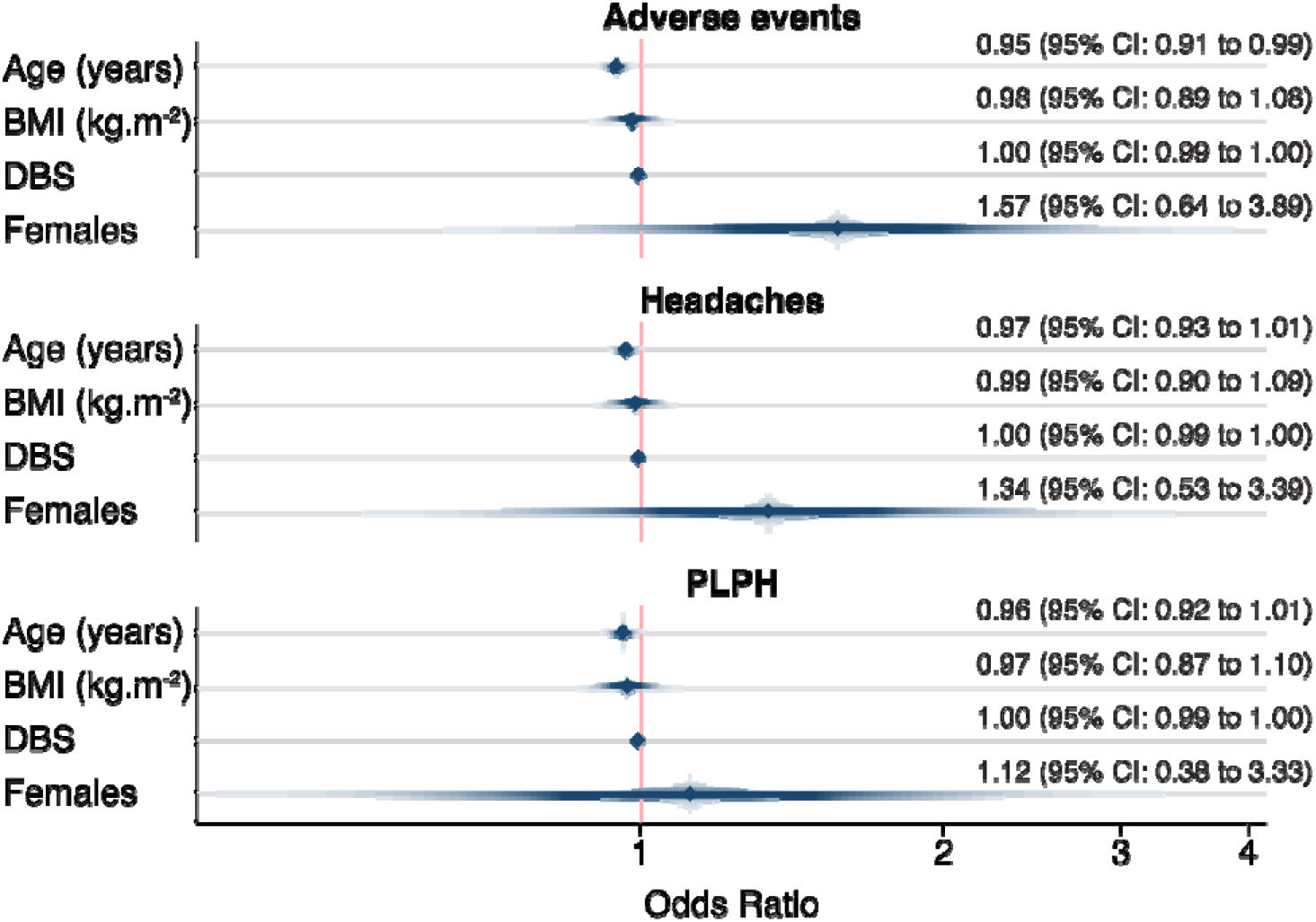
Adverse events’ association with age, gender, BMI and DBS in gene expansion carriers. PLPH, post-lumbar puncture headache; BMI, body mass index; DBS, Disease Burden Score.

### CSF quality

10,610 mL of CSF was collected, of which 10,430.5 mL (98.31%) were deemed usable, amounting to 32,808 300μL-cryovials of CSF (Figure 3). Each LP produced a median of 20mL of CSF (IQR 0, minimum 2, maximum 24), of which a median of 20mL were deemed usable (IQR 1, minimum 2, maximum 24), generating a median of 63 cryovials of CSF (IQR 11, minimum 1, maximum 86). CSF sample processing began a median of 6 min from the end of collection (IQR 7.5, minimum 1, maximum 51); the processing itself took a median of 27 min (IQR 11, minimum 10, maximum 132), and samples were stored in the freezer in a further 1 min (IQR 2, minimum 0, maximum 242). Overall the median time from collection to storage was 36 minutes (IQR 15, minimum 12, maximum 292).

**Figure 3.**
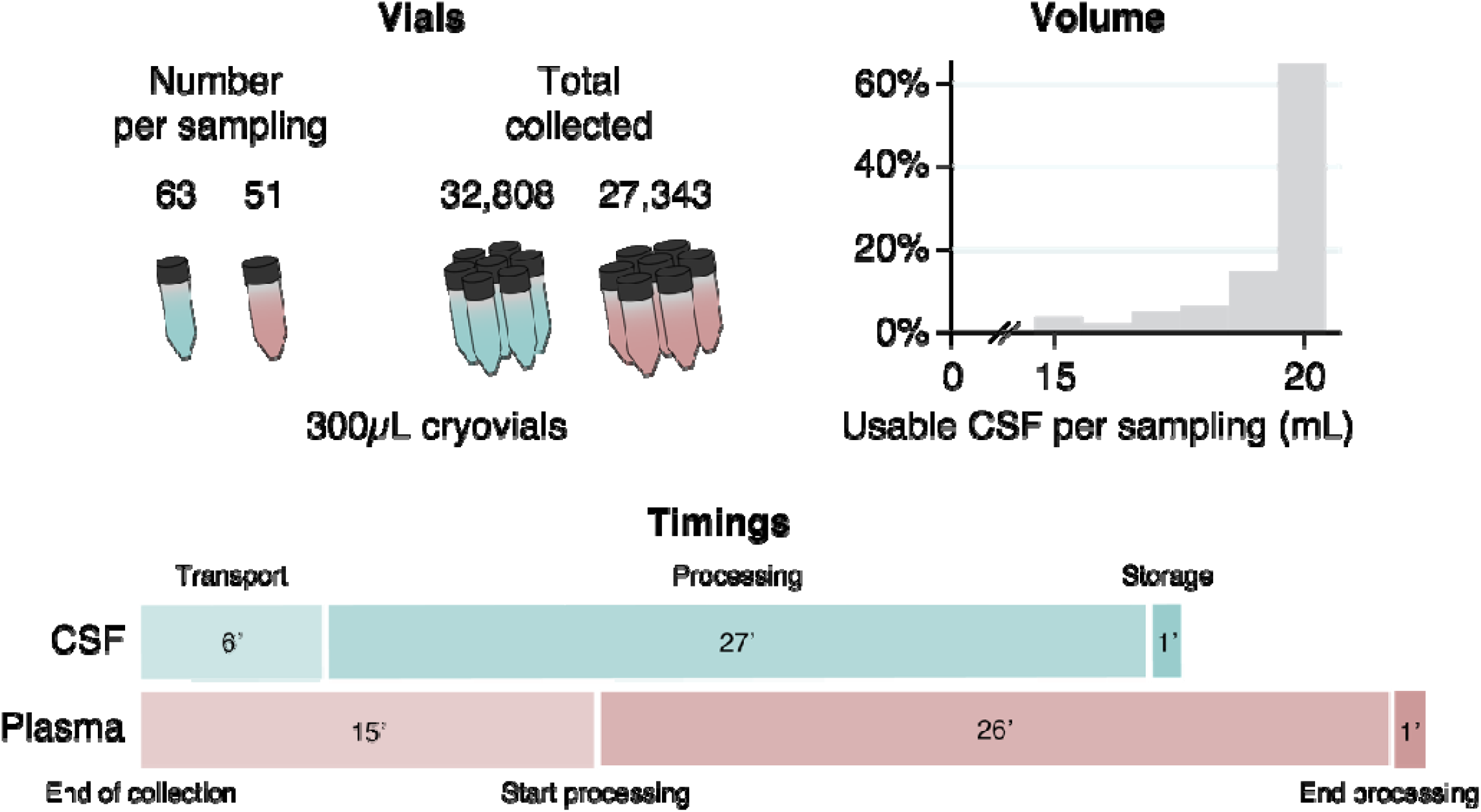
Number of vials collected, volume of usable CSF per sampling visit, and median biosample processing times in minutes.

Samples had a median of 0 white blood cells per μL (IQR 1, minimum 0, maximum 24) and 1 red blood cell per μL (IQR 6, minimum 0, maximum 2,645). Haemoglobin was measured in 245 samples and the median concentration was 0.322 μg/ml (IQR 0.731, minimum 0, maximum 17.080). 224 (91.43%) were below 2 μg/ml, defined as the maximum acceptable amount of blood contamination for accurate quantification of mutant huntingtin (one critical measurement in HD research) by the most commonly used assay(30).

### Blood product quality

So far 27,343 300μL-cryovials of 300μL plasma (Figure 3) and 1,142 1500μL-cryovials of serum have been collected. Each participant donated a median of 51 cryovials of plasma (IQR 17, minimum 2, maximum 75) and 2 cryovials of serum (IQR 0, minimum 1, maximum 3). Overall the median time from blood collection to storage was 47 minutes (IQR 19, minimum 14 maximum 288). Each plasma sample took a median of 15 min from the end of collection to start processing (IQR 12, minimum 1, maximum 150), the processing took a median of 26 min (IQR 13, minimum 10, maximum 93), and samples were stored in the freezer in 1 min (IQR 5, minimum 0, maximum 217). Each serum sample took a median of 17 min from the end of collection to start processing (IQR 20, minimum 1, maximum 169), the processing took a median of 14 min (IQR 7.5, minimum 10, maximum 66), and samples were stored in the freezer in 8 min (IQR 17, minimum 0, maximum 189)

### Sample distribution

There have been 14 sample requests from qualified investigators (6 from industry and 8 from academia or non-profit organizations), of which 11 have been approved by an independent scientific review committee, and 3 are currently being evaluated. In total 3,251 CSF cryovials, 2,662 plasma cryovials and 238 serum cryovials have been shipped to collaborators.

## DISCUSSION

The HDClarity study is the largest CSF collection initiative in HD, with the added advantage of matched phenotyping and plasma/serum samples in both, HDGECs and healthy controls. The experience reported here of the first 572 sampling visits over 43 months, with low rates of screen failure (2%) and adverse events (24.13%) suggests that LPs in HD have a safety profile akin to what is known in the healthy population. Large-scale, international, multisite biosample collection initiatives including CSF collection are viable in the HD population across all disease stages(31).

Having phenotypic data matched for all CSF samples will facilitate validation of future biomarkers while blood will grant the opportunity to explore correlations between blood and CSF levels of biomarkers. Over 100 repeat sampling visits, conducted 4 to 8 weeks after initial sampling visit permit the study of the short-term stability of potential biomarkers – crucial for the generation of sample size calculations and longitudinal clinical study designs(8).

Adverse event rates in our study are aligned with what has been reported in literature for HD(10), Parkinson’s(32) and Alzheimer’s disease(33-35). In a multivariable analysis, we did not find any factors such as gender, age and BMI to influence the frequency of adverse events, including PLPH. DBS, as a continuous surrogate of disease state, did not seem to associate with the frequency of headaches, although participants with manifest HD had fewer events than healthy controls. In the only serious adverse event, where the subject underwent a blood patch for relief of PLPH, the procedure had been performed using a larger-bore, LP needle with a cutting tip, provided locally at the site rather than from the HDClarity kit. Causation between use of a different needle and this PLPH cannot be established. The study uses 22G pencil-point (Whitacre) needles and will shortly switch to 24G needles which may be helpful in reducing adverse events of back pain and headache. Lack of standardisation in methodology, which has been a frequent limitation of biomarker research in HD(13) ought to be much less of an issue, especially since the HDClarity protocol is open-access.

The data from HDClarity show that LP(s) is generally safe and tolerable in HDGECs. This will allow wide scale exploration of future biomarker avenues. Currently proposed biomarker candidates of HD disease progression, such as neurofilament light (NFL) and mutant huntingtin (mHTT), need to be validated in larger longitudinal cohorts(8, 14). The combination of HDClarity samples and Enroll-HD data provide the tool for testing and validating biomarkers in HD research and drug development, while the Enroll-HD platform has greatly assisted in recruitment of sites and participants. Studies proposed using the HDClarity biorepository aim at studying the utility of candidate biomarkers (e.g. mHTT, total HTT, NFL, YKL-40, total Tau, phospho-Tau, IL6) in predicting progression through different stages of HD with an emphasis on premanifest and early HD. Cross sectional studies will be used to identify most promising candidates, other than mHTT and NFL, followed by longitudinal validation studies. The current recruitment goal of 1,200 participants balanced across study groups will enable such analyses incorporating sophisticated modelling of data accounting for confounding factors. Eventually, a high-quality representative data set will be generated depicting the profile of each biochemical biomarker that can be utilized by all stakeholders for research and drug development. ImageClarity, another prospective nested study, will be launched in the near future where HDClarity participants who are willing and eligible will undergo a multisequence brain MRI that incorporates structural and functional modalities-offering a valuable opportunity to unite clinical, biofluid and imaging data for each participant. Availability of longitudinal phenotypic, biosample and imaging data on HDGECS at all stages and healthy controls will be a powerful asset in biomarker development for HD. Interested investigators should visit http://hdclarity.net for more information.

## Data Availability

The datasets generated during and/or analysed during the current study are available from the corresponding author on reasonable request.

## Acknowledgements

We would like to thank all the participants from the HD community who donated samples and gave their time to take part in this study, Robi Blumenstein for his support in the development and implementation of HDClarity, and Stefanie Gosling, Mette Gilling, Olivia Handley and Eileen Neacy for their contributions to the conduct of the project. Samples and data used in this work were generously provided by the participants in the HDClarity and HD-CSF studies and made available by CHDI Foundation, Inc.

Data used in this work were generously provided by the participants in the Enroll-HD study and made available by CHDI Foundation, Inc. Enroll-HD is a global clinical research platform intended to accelerate progress towards therapeutics for Huntington’s disease; core datasets are collected annually on all research participants as part of this multi-center longitudinal observational study. Enroll-HD is sponsored by CHDI Foundation, Inc., a nonprofit biomedical research organization exclusively dedicated to developing therapeutics for Huntington’s disease. Enroll-HD would not be possible without the vital contribution of the research participants and their families.

Samples and data used in this work would not be possible without the vital contribution of the research participants and their families in the HDClarity and HD-CSF studies. HDClarity and HD-CSF are cerebrospinal fluid collection initiatives designed to facilitate therapeutic development for Huntington’s disease. HDClarity and HD-CSF are led by Dr. Edward Wild and sponsored by University College London. HDClarity is funded by CHDI Foundation, Inc., a nonprofit biomedical research organization exclusively dedicated to developing therapeutics that will substantially improve the lives of those affected by Huntington’s disease. The Medical Research Council UK (MR/M008592/1) funded HD-CSF.

The HDClarity Investigators are Edward J Wild, Mark Guttman, Blair Roland Leavitt, Ralf Reilmann, Carsten Saft, Jürgen Winkler, Zacharias Kohl, Jan Lewerenz, Hugh Rickards, Stuart Ritchie, Jeremy Cosgrove, Nayana Lahiri, Roger Barker, Jee Bang, Francis Walker, Erin Furr-Stimming (more details in Supplementary Appendix 3).

## Author’s Roles

EJW and CS designed the study. FBR, BRL, MG, JB and Jlew were involved in participant recruitment, eligibility check, clinical examinations and sample collection. DK and EP managed sample storage. GO and KS monitored data. GO, EP, SL, JT, KS and Jlev managed the study. FBR developed and performed the statistical analysis; FBR and EJW interpreted the data and wrote the manuscript; and all authors contributed to reviewing the manuscript.

## Tables

**Table 3.**
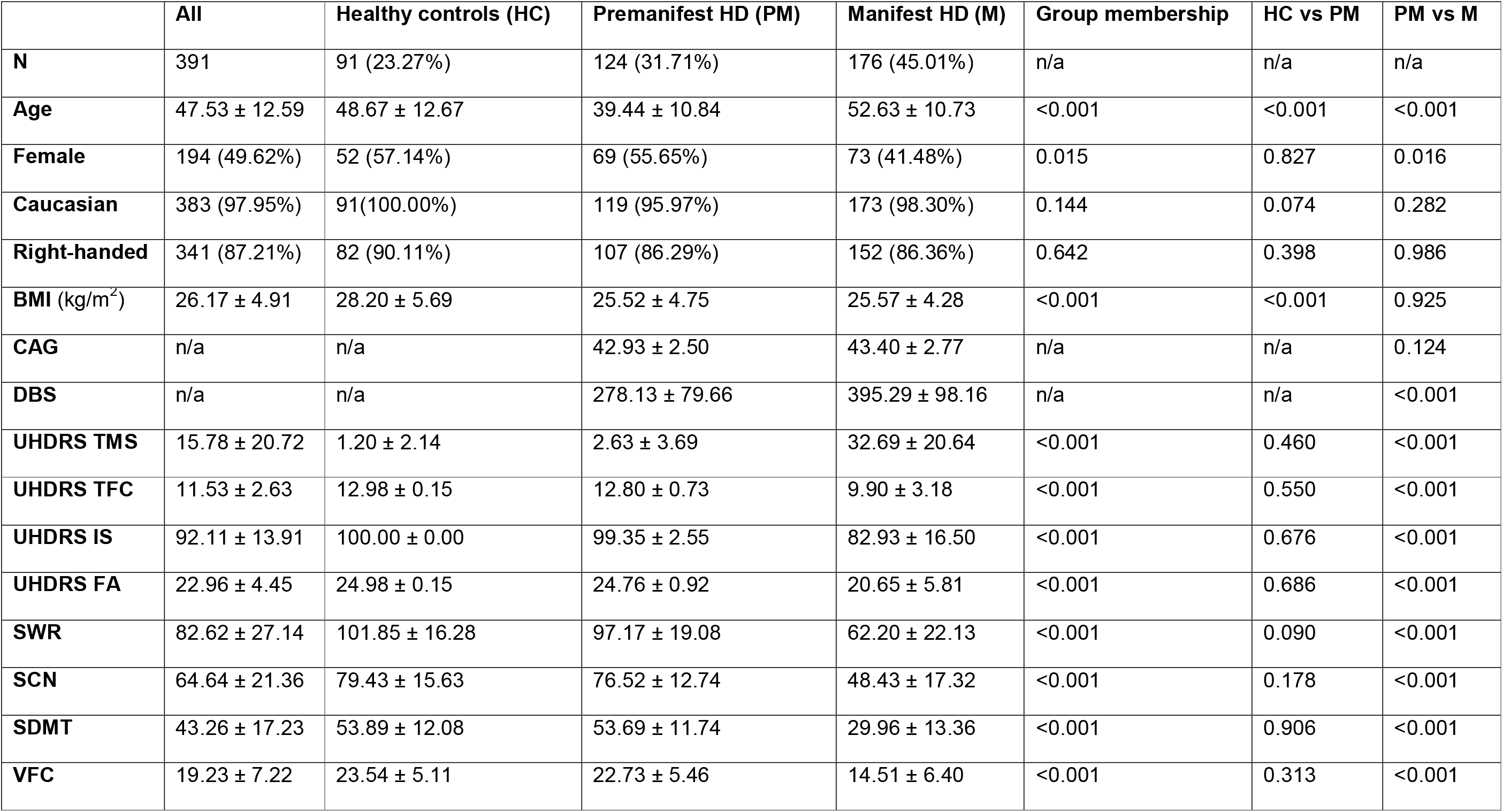
Characteristics at first screening visit of participants who underwent at least 1 successful sampling visit. Successful visits were defined as when dura was pierced and CSF was collected, irrespective of amount of CSF. Continuous variables are reported as mean ± standard deviations. Categorical variables are reported as absolute and relative frequencies. BMI, body mass index; CAG, CAG repeat count; DBS, Disease Burden Score; UHDRS, Unified Huntington’s Disease Rating Scale; TMS, UHDRS Total Motor Score; TFC, UHDRS Total Functional Capacity; IS, UHDRS Independence Score; FA, UHDRS Functional Assessment; SWR, Stroop Word Reading test; SCN, Stroop Color Naming test; SDMT, Symbol Digits Modality Test; VFC, Verbal Fluency Categorical; n/a, not applicable.

|  | All | Healthy controls (HC) | Premanifest HD (PM) | Manifest HD (M) | Group membership | HC vs PM | PM vs M |
| --- | --- | --- | --- | --- | --- | --- | --- |
| **N** | 391 | 91 (23.27%) | 124 (31.71%) | 176 (45.01%) | n/a | n/a | n/a |
| **Age** | 47.53 ± 12.59 | 48.67 ± 12.67 | 39.44 ± 10.84 | 52.63 ± 10.73 | <0.001 | <0.001 | <0.001 |
| **Female** | 194 (49.62%) | 52 (57.14%) | 69 (55.65%) | 73 (41.48%) | 0.015 | 0.827 | 0.016 |
| **Caucasian** | 383 (97.95%) | 91(100.00%) | 119 (95.97%) | 173 (98.30%) | 0.144 | 0.074 | 0.282 |
| **Right-handed** | 341 (87.21%) | 82 (90.11%) | 107 (86.29%) | 152 (86.36%) | 0.642 | 0.398 | 0.986 |
| **BMI (kg/m<sup>2</sup>)** | 26.17 ± 4.91 | 28.20 ± 5.69 | 25.52 ± 4.75 | 25.57 ± 4.28 | <0.001 | <0.001 | 0.925 |
| **CAG** | n/a | n/a | 42.93 ± 2.50 | 43.40 ± 2.77 | n/a | n/a | 0.124 |
| **DBS** | n/a | n/a | 278.13 ± 79.66 | 395.29 ± 98.16 | n/a | n/a | <0.001 |
| **UHDRS TMS** | 15.78 ± 20.72 | 1.20 ± 2.14 | 2.63 ± 3.69 | 32.69 ± 20.64 | <0.001 | 0.460 | <0.001 |
| **UHDRS TFC** | 11.53 ± 2.63 | 12.98 ± 0.15 | 12.80 ± 0.73 | 9.90 ± 3.18 | <0.001 | 0.550 | <0.001 |
| **UHDRS IS** | 92.11 ± 13.91 | 100.00 ± 0.00 | 99.35 ± 2.55 | 82.93 ± 16.50 | <0.001 | 0.676 | <0.001 |
| **UHDRS FA** | 22.96 ± 4.45 | 24.98 ± 0.15 | 24.76 ± 0.92 | 20.65 ± 5.81 | <0.001 | 0.686 | <0.001 |
| **SWR** | 82.62 ± 27.14 | 101.85 ± 16.28 | 97.17 ± 19.08 | 62.20 ± 22.13 | <0.001 | 0.090 | <0.001 |
| **SCN** | 64.64 ± 21.36 | 79.43 ± 15.63 | 76.52 ± 12.74 | 48.43 ± 17.32 | <0.001 | 0.178 | <0.001 |
| **SDMT** | 43.26 ± 17.23 | 53.89 ± 12.08 | 53.69 ± 11.74 | 29.96 ± 13.36 | <0.001 | 0.906 | <0.001 |
| **VFC** | 19.23 ± 7.22 | 23.54 ± 5.11 | 22.73 ± 5.46 | 14.51 ± 6.40 | <0.001 | 0.313 | <0.001 |

**Table 4.** Frequency of adverse events per visit. Categorical variables are reported as absolute and relative frequencies. PLPH, post-lumbar puncture headache; HD, Huntington’s disease

|  | Absolute and relative frequencies |  |  |  | Group membership | HC vs PM | HC vs M |
| --- | --- | --- | --- | --- | --- | --- | --- |
|  | All | Healthy controls<br>(HC) | Premanifest HD<br>(PM) | Manifest HD<br>(M) | p-value | p-value | p-value |
| **Adverse events** | 138 (24.13%) | 34 (26.15%) | 55 (31.07%) | 49 (18.49%) | 0.432 | 0.633 | 0.205 |
| **Headaches** | 85 (14.86%) | 26 (20.00%) | 32 (18.08%) | 27 (10.19%) | 0.065 | 0.305 | 0.020 |
| **PLPH** | 70 (12.24%) | 22 (16.92%) | 26 (14.69%) | 22 (8.30%) | 0.099 | 0.195 | 0.033 |

