## Supplementary materials for "Safety and feasibility of research lumbar puncture in Huntington’s disease: the HDClarity cohort and bioresource"

**Word count:** 3,484
**Abstract:** 250
**Tables/Illustrations:** 5

**References:** 40

### SUPPLEMENTARTY MATERIAL

#### Supplementary figures

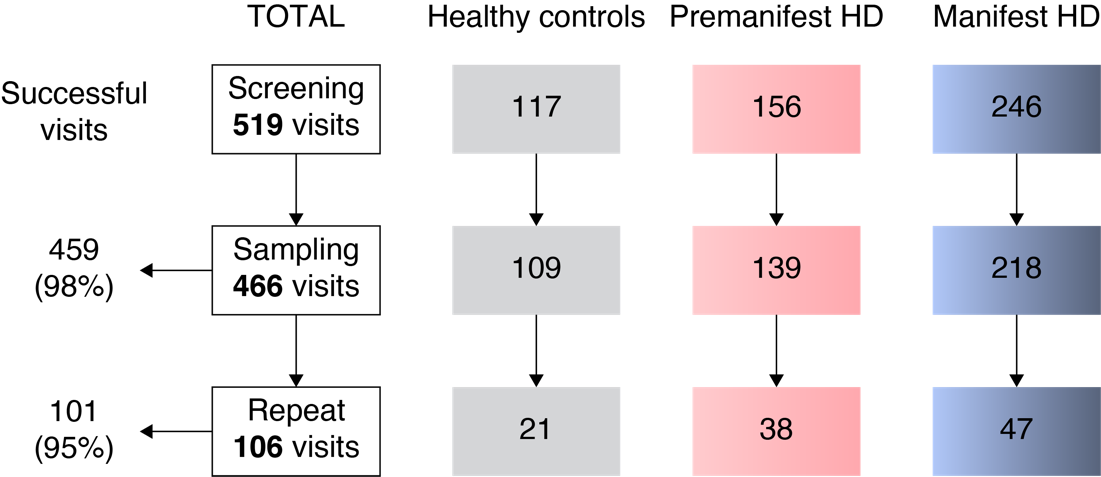

**Figure S1-** **Visit disposition from screening to sampling and short-term repeat sampling visit, by disease group.** Successful visits were defined as when dura was pierced and CSF was collected, irrespective of amount of CSF. HD, Huntington’s disease.

#### Supplementary tables

**Table S1 – Participant cohorts**

| **Cohort** | | **CAG** | **DBS** | **UHDRS DCS** | **UHDRS TFC** |
| --- | --- | --- | --- | --- | --- |
| Healthy controls | | <36  (or no known family history) | - | - | - |
| Gene expansion carriers | Early premanifest | ≥ 40 | < 250 | < 4 | - |
|  | Late premanifest | ≥ 40 | ≥ 250 | < 4 | - |
|  | Early manifest | ≥ 36* | - | 4 | 7-13 |
|  | Moderate manifest | ≥ 36* | - | 4 | 4-6 |
|  | Late manifest | ≥ 36* | - | 4 | 0-2 |

*the current protocol (version 3 from 19 December 2018) requires manifest gene expansion carriers to have a CAG repeat count of 40 or more, but previous versions allowed participants with ≥ 36 repeats (version 1 from 6 October 2015, and 2 from 21 June 2016). CAG, CAG repeat count; DBS, Disease Burden Score ((CAG – 35.5) × age); DCS, Diagnostic Confidence Score; TFC, Total Functional Capacity; UHDRS, Unified Huntington’s Disease Rating Scale.

**Table S2 – Comparison of participants characteristics at successful (i.e. when dura was pierced and CSF was collected, irrespective of amount of CSF) and unsuccessful visits (i.e. where an LP was attempted, but no CSF was collected).**

|  | **Successful visit** | **Unsuccessful visit** | **p-value** |
| --- | --- | --- | --- |
| **N** | 560 (97.90%) | 12 (2.10%) | n/a |
| **Age** | 48.35 ± 12.63 | 52.42 ± 9.97 | 0.272 |
| **Female** | 272 (48.57%) | 7 (58.33%) | 0.506 |
| **Caucasian** | 549 (98.04%) | 12 (100.00%) | 0.624 |
| **Right-handed** | 488 (87.14%) | 10 (83.33%) | 0.404 |
| **BMI** (kg/m^2^) | 26.09 ± 5.00 | 29.94 ± 5.80 | 0.064 |
| **Study Cohort** | **HC:** 129 (23.04%)  **PM:** 259 (46.25%)  **M**: 172 (30.71%) | **HC:** 1 (8.33%)  **PM:** 5 (50.00%)  **M**: 6 (41.67%) | 0.487 |
| **UHDRS TMS** | 17.19 ± 22.30 | 24.67 ± 29.57 | 0.260 |
| **UHDRS TFC** | 11.33 ± 2.88 | 10.92 ± 3.45 | 0.624 |
| **UHDRS IS** | 90.83 ± 15.43 | 87.50 ± 20.17 | 0.465 |
| **UHDRS FA** | 22.55 ± 4.94 | 21.33 ± 6.85 | 0.406 |
| **SWR** | 81.88 ± 28.60 | 67.75 ± 25.36 | 0.095 |
| **SCN** | 64.32 ± 22.40 | 49.33 ± 15.50 | 0.108 |
| **SDMT** | 42.94 ± 17.93 | 32.75 ± 14.38 | 0.175 |
| **VFC** | 19.12 ± 7.36 | 16.25 ± 5.63 | 0.183 |

*Continuous variables are reported as mean ± standard deviations. Categorical variables are reported as absolute and relative frequencies. BMI, body mass index; HC, healthy controls; PM, premanifest HD; M, manifest HD; UHDRS, Unified Huntington’s Disease Rating Scale; TMS, UHDRS Total Motor Score; TFC, UHDRS Total Functional Capacity; IS, UHDRS Independence Score; FA, UHDRS Functional Assessment; SWR, Stroop Word Reading test; SCN, Stroop Color Naming test; SDMT, Symbol Digits Modality Test; VFC, Verbal Fluency Categorical; n/a, not applicable.*

**Table S3 – Comparison of participants characteristics at visits with and without adverse events, headaches, and post-lumbar puncture headache**

|  | **Adverse events** | | | **Headaches** | | | | **Post-lumbar puncture headache** | | |
| --- | --- | --- | --- | --- | --- | --- | --- | --- | --- | --- |
|  | **No** | **Yes** | **p-value** | **No** | **Yes** | **p-value** | **No** | | **Yes** | **p-value** |
| **N** | 434 (75.87%) | 138 (24.13%) | n/a | 487 (85.14%) | 85 (14.86%) | n/a | 502 (87.76%) | | 70 (12.24%) | n/a |
| **Age** | 49.84 ± 12.42 | 44.01 ± 12.09 | >0.001 | 48.96 ± 12.60 | 45.43 ± 12.16 | 0.024 | 49.00 ± 12.53 | | 44.42 ± 12.34 | 0.008 |
| **Female** | 200 (46.08 %) | 79 (57.25%) | 0.048 | 229 (47.02%) | 50 (58.82%) | 0.057 | 239 (47.61%) | | 40 (57.14%) | 0.155 |
| **Caucasian** | 427 (98.39%) | 134 (97.10%) | 0.402 | 479 (98.36%) | 82 (96.47423%) | 0.313 | 492 (98.01%) | | 69 (98.57%) | 0.793 |
| **Right-handed** | 375 (86.41%) | 123 (89.13%) | 0.297 | 423 (86.86%) | 75 (88.24%) | 0.428 | 435 (86.65%) | | 63 (90.00%) | 0.267 |
| **BMI** (kg/m^2^) | 26.36 ± 5.22 | 25.57 ± 4.41 | 0.113 | 26.28 ± 5.13 | 25.55 ± 4.46 | 0.233 | 26.27 ± 5.12 | | 25.42 ± 4.36 | 0.213 |
| **Study Cohort** | **HC:** 96 (22.12%)  **PM:** 122 (28.11%)  **M**: 216 (49.77%) | **HC:** 34 (24.64%)  **PM:** 55 (39.86%)  **M**: 49 (35.51%) | 0.023 | **HC:** 104 (21.36%)  **PM:** 145 (29.77%)  **M**: 238 (48.87%) | **HC:** 26 (30.59%)  **PM:** 32 (37.65%)  **M**: 27 (31.76%) | 0.023 | **HC:** 108 (21.51%)  **PM:** 151 (30.08%)  **M**: 243 (48.41%) | | **HC:** 22 (31.43%)  **PM:** 26 (37.14%)  **M**: 22 (31.43%) | 0.041 |
| **UHDRS TMS** | 19.68 ± 24.08 | 10.04 ± 14.19 | >0.001 | 18.83 ± 23.52 | 8.88 ± 12.15 | 0.001 | 18.61 ± 23.31 | | 8.34 ± 11.79 | 0.001 |
| **UHDRS TFC** | 11.08 ± 3.10 | 12.08 ± 1.91 | 0.002 | 11.17 ± 3.03 | 12.19 ± 1.60 | 0.006 | 11.20 ± 3.00 | | 12.21 ± 1.61 | 0.011 |
| **UHDRS IS** | 89.48 ± 16.59 | 94.78 ± 10.65 | 0.002 | 89.89 ± 16.36 | 95.76 ± 7.73 | 0.003 | 90.10 ± 16.22 | | 95.50 ± 7.72 | 0.012 |
| **UHDRS FA** | 22.10 ± 5.41 | 23.88 ± 2.92 | 0.002 | 22.25 ± 5.29 | 24.14 ± 1.85 | 0.004 | 22.30 ± 5.24 | | 24.19 ± 1.76 | 0.010 |
| **SWR** | 79.35 ± 29.41 | 88.52 ± 24.71 | 0.004 | 79.99 ± 28.98 | 90.61 ± 24.50 | 0.004 | 80.08 ± 29.00 | | 92.26 ± 22.97 | 0.003 |
| **SCN** | 62.39 ± 22.96 | 69.01 ± 19.70 | 0.007 | 62.84 ± 22.66 | 70.59 ± 19.55 | 0.006 | 62.88 ± 22.72 | | 71.96 ± 17.94 | 0.004 |
| **SDMT** | 40.89 ± 18.03 | 48.35 ± 16.39 | >0.001 | 41.57 ± 17.90 | 49.16 ± 16.66 | 0.001 | 41.68 ± 17.95 | | 50.03 ± 16.00 | 0.001 |
| **VFC** | 18.44 ± 7.52 | 21.00 ± 6.39 | 0.002 | 18.75 ± 7.51 | 20.82 ± 6.02 | 0.025 | 18.78 ± 7.49 | | 21.06 ± 5.84 | 0.025 |

*Continuous variables are reported as mean ± standard deviations. Categorical variables are reported as absolute and relative frequencies. BMI, body mass index; HC, healthy controls; PM, premanifest HD; M, manifest HD; UHDRS, Unified Huntington’s Disease Rating Scale; TMS, UHDRS Total Motor Score; TFC, UHDRS Total Functional Capacity; IS, UHDRS Independence Score; FA, UHDRS Functional Assessment; SWR, Stroop Word Reading test; SCN, Stroop Color Naming test; SDMT, Symbol Digits Modality Test; VFC, Verbal Fluency Categorical; n/a, not applicable.*

**Table S4 –** **Frequency of adverse events.** Categorical variables are reported as absolute and relative frequencies.

| **Absolute and relative frequencies (n=134)** | | |
| --- | --- | --- |
| Headache | 90 | 47.62% |
| Back pain | 54 | 28.57% |
| Vasovagal reactions | 10 | 05.29% |
| Nausea & vomiting | 7 | 03.70% |
| Bruising | 7 | 03.70% |
| Paraesthesia | 2 | 01.06% |
| Other | 19 | 10.05% |

#### Supplementary appendix 1 - Ethics Committees / Institutional Review Boards decisions

*Center Movement Disorders, CA:*

- Approval by Western Institutional Review Board (STUDY NUM: 1169046)

*University British Columbia, CA:*

- Approval by Clinical Research Ethics Board, University of British Columbia (UBC CREB NUMBER: H16-02183)

*George Huntington Institute, DE:*

- Approval by Ethik Kommission der Ärztekammer Westfalen-Lippe und der Westfälischen Wilhelms-Universität (Unser Aktenzeichen: 2017-130-b-S)

*St Josef And Elisabeth Hospital, DE:*

- Approval by Ethik-Kommission der Medizinischen Fakultät der Ruhr Universität Bochum (Reg.-Nr.: 17-5997)

*University Hospital Erlangen, DE:*

- Approval by Ethik-Kommission der Friedrich-Alexander Universität (Antrag Nr.: 285_17 Bc)

*University Hospital Ulm, DE:*

- Approval by Ethikkommission der Universität Ulm (Antrag Nr. 286/16)

*BirmSolNHSFounTrust, UK:*

- UK-wide approval by London – Camberwell St Giles Research Ethics Committee (REC reference: 16/LO/1878)
- Local approval by Birmingham and Solihull Mental Health NHS Foundation Trust (IRAS 185506)

*GreatGlasgowHealthBoard, UK:*

- UK-wide approval by London – Camberwell St Giles Research Ethics Committee (REC reference: 16/LO/1878)
- Local approval by Greater Glasgow and Clyde Health Board (R&D reference: GN16NE605)

*LeedsTeachHospTrust, UK:*

- UK-wide approval by London – Camberwell St Giles Research Ethics Committee (REC reference: 16/LO/1878)
- Local approval by Leeds Teaching Hospitals NHS Trust (R&I No: CG16/81309)

StGeorgeHealthTrust, *UK*:

- UK-wide approval by London – Camberwell St Giles Research Ethics Committee (REC reference: 16/LO/1878)
- Local approval by St George's University Hospitals NHS Foundation Trust (R&D number 17.0054)

University Cambridge, *UK*:

- UK-wide approval by London – Camberwell St Giles Research Ethics Committee (REC reference: 16/LO/1878)
- Local approval by Research and Development Department, Cambridge University Hospitals NHS Foundation Trust (R&D ref: A094293)

*University College London, UK:*

- UK-wide approval by London – Camberwell St Giles Research Ethics Committee (REC reference: 16/LO/1878)
- Local approval by London – Camberwell St Giles Research Ethics Committee (REC reference: 16/LO/1917)

*JohnsHopkinsUniv, US:*

- Approval by Office of Human Subject Research Institutional Review Boards, Johns Hopkins (Number: IRB00104965)

*WakeForestUniv, US:*

- Approval by Wake Forest University School of Medicine Institutional Review Board (IRB Number: IRB00040219)

*UnivTexasHlthCntrHous, US:*

- Approval by Committee for the Protection of Human Subjects, UTHealth, the University of Texas (HSC-MS-17-0731)

#### Supplementary appendix 2 - Biosample collection and processing procedures

##### CSF collection

**LUMBAR PUNCTURE**

1. Identify L4/5 or L3/4 space using surface markings (i.e. the intercristal line)
2. Place subject into lateral decubitus position with pillow between knees
3. Disinfect skin using antiseptic applicator.
4. It is highly recommended to use adequate lidocaine to reduce the discomfort of this LP procedure. If, after noting allergies or sensitivities to lidocaine and discussing the risks and benefits of local anaesthesia, it is decided to forgo this step, it should be noted in the case report form. Inject up to 5ml of 2% lidocaine for local anaesthesia. Use the 25G 1" needle and inject lidocaine to raise a skin wheal. Then inject lidocaine more deeply using the 21G needle.
5. Obtain CSF using the supplied spinal needle. If the participant is thin, do not insert the deep infiltration needle all the way. Use only about 2/3 of its length (to prevent entering the subarachnoid space with anything other than the pencil-point spinal needle).
6. If CSF cannot be obtained, up to three needles may be used. An alternative design of spinal needle supplied by the site may be used if, after at least one attempt with the supplied needle, it is felt this will increase the chance of success.
7. If the CSF collection fails then there is no need to collect blood samples from the participant at this visit
8. An adjacent space may be used (with further lidocaine, max. total 10 ml, if needed).
9. If necessary, the CSF space may be located by sitting participant up, but once CSF is seen, it is recommended to have participant lie back in lateral decubitus position for 30 seconds before collection begins. Document positions of participant during puncture and collection in the eCRF
10. Document the space used for lumbar puncture, the number of needle passes (i.e. the number of times a needle is inserted and removed from the skin), the number of attempts (i.e. the number of times the lumbar space, the participant position, or the investigator conducting the LP change), the volume of lidocaine used, and the time CSF collection started and ended in the eCRF
11. Omit pressure measurement for all subjects (this is because polypropylene manometers are not available)
12. CSF is collected without suction in 50ml tubes placed on wet ice in the Styrofoam cup
13. Collect the first 1 ml of CSF into the supplied tube labelled ‘CSF’. If the first 1 ml (approx. 15 drops) is not macroscopically bloody, continue sampling CSF in the same tube up to 15-20 ml, as allowed locally, keeping the tube in the wet ice cup.

    If the first 1 ml is macroscopically bloody,

- stop collecting CSF by reinserting the stylet partially
- Discard the tube, and collect a second 1 ml in a new pre-cooled ‘CSF’ tube, and examine it visually for blood contamination
- If it is free of blood, continue collecting CSF up to 14-19 ml (1ml less than the locally permitted maximum).
- If the second separately collected ml of CSF is also macroscopically bloody, discard the tube, and continue to collect 13-18 ml of CSF in a third pre-cooled ‘CSF’ tube.
- If the third tube is macroscopically bloody, stop collecting and abandon the procedure or attempt the LP in a different space, if there is reason to believe blood-free CSF can be obtained. You may need to open a new collection kit to provide sufficient tubes; if this creates any discrepancies in the kit ID numbers, it must be noted carefully and explained in the eCRF.
- Stop collecting CSF when sampling time exceeds 20 minutes. Document these details in the eCRF.

14. Place cap on tube and leave on wet ice until further processing.
15. Reinsert the stylet before withdrawing the needle.
16. Cover the puncture site with sterile dressing.
17. Record time of CSF collection (time when CSF was first seen).
18. At the discretion of the Site Principal Investigator, participants may be instructed to lie flat for 1 hour.

Transport CSF immediately to laboratory for processing, do not wait for the blood samples to be ready as this can cause delays

##### Blood collection

| ! Please make sure that all caps are tightly secured.  ! Check the expiration date on the tube- do not use expired tubes!  ! Do not collect blood samples if CSF collection was not successful! | |
| --- | --- |
| Specimens are best collected through venipuncture using a butterfly needle vacuumed directly into the required tube. | 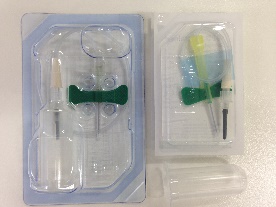 |
| 1. Fill 4 x 10 ml blood in lithium heparin tubes | 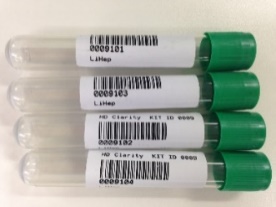 |
| 2. Gently invert each lithium heparin tube 10 times immediately after collection, and place on wet ice |  |
| 3. Fill 1 8.5ml serum tube | 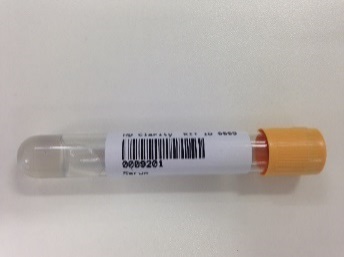 |
| 4. Immediately after collection transfer all blood samples to the lab for processing |  |

##### CSF processing

| **Sample Collection** | 1. Lab to receive one 50ml CSF collection tube filled up to 20mls with CSF   (collected from participant between 08:00 - 10:30 local time)  4 tubes are provided in case of blood contamination. All clean CSF sent to the lab should be in a single tube. | 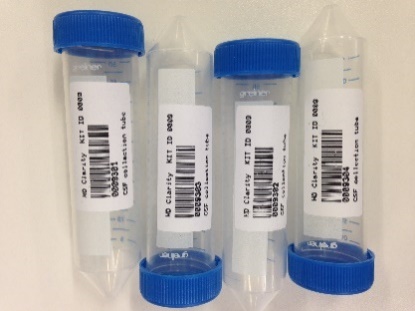 |
| --- | --- | --- |
|  | 2. CSF sample is collected while the collection tube is in the styrofoam cup filled with wet ice.   Sample is transported to the lab in wet ice (container to be supplied by site). | 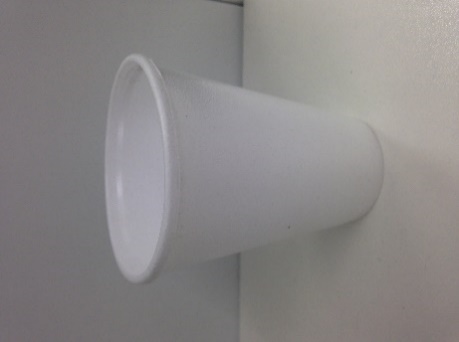 |
|  | 3. Samples transported immediately to laboratory for processing. | 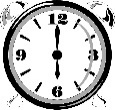  **Processing must start within 15 minutes of sample collection** |
|  | 4. After CSF collection, details including the Kit ID are recorded in the **CSF** eCRF, ‘CSF collection’ box (Screen shot 2). |  |
| **Sample Processing** | 5. Note the CSF processing start time |  |
|  | 6. Agitate the entire CSF sample for 10 seconds using a vortex mixer to homogenise CSF |  |
|  | 7. Using a sterile individually wrapped polypropylene 1ml pipette tip, extract 200 µl of the CSF and use it to determine white blood cell count and erythrocyte count per μl in triplicate according to local GLP- approved laboratory practice as instructed at the Site initiation visit and in the Manual CSF Cell Count SOP   Cell counts should be recorded on the ‘CSF Quality’ eCRF in the ‘Onsite CSF Sample Quality Control’ box (Screen shot 3) | 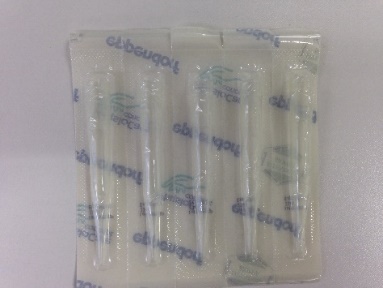  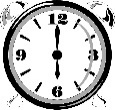  **Triplicate cell count should be done within 60 minutes of sample collection.** |
|  | 8. Balance the centrifuge and before filling the balance tube with water please clearly mark the tube so that it can easily be identified as water (not CSF). 9. Centrifuge the 50ml tube containing residual CSF at **400 × g for 10 min at 4°C** to remove cells while preserving cell integrity for potential future use. Cell integrety in needed so that intracelular substances do not contaminate the non-cellular phase of the CSF | **Label your balance tube!** |
|  | 10. Using the polypropylene Pasteur pipette, transfer the supernatant into a single 30ml polypropylene tube labelled “CSF supernatant” and agitate for 10 seconds to homogenise CSF   **If the polypropylene Pasteur pipettes are damaged then it is acceptable to decant the supernatant into the tube. No pipettes should be used other than those supplied.** | 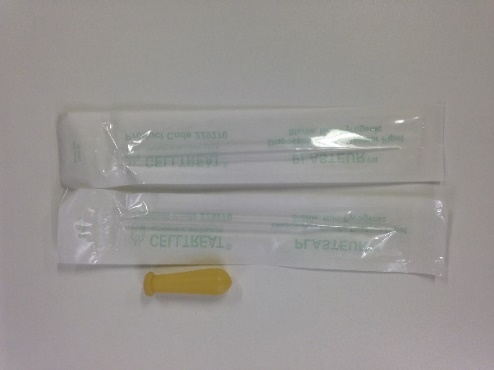  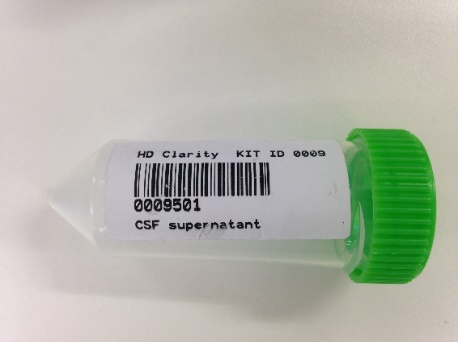 |
|  | 11. Aliquot the CSF in 300 µl aliquots into the cryovials labelled “CSF”, using a sterile individually wrapped polypropylene 1ml pipette tip   Note the tube rack ID, tube ID (this must be the same for all aliquots) and the number of aliquots for later recording on the eCRF. Please dispose of any unused aliquots  CSF aliquots must have blue lids. Any samples that do not have the expected lid colour will be discarded by BioRep. | 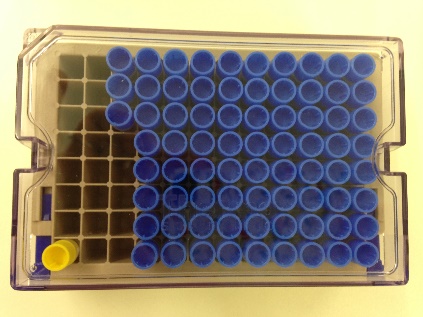 |
|  | 12. Re-suspend the CSF cell pellet in 300 µl of supplied RNA*later* solution, using gentle vortex agitation, and use another sterile pipette tip to transfer to a cryovial with yellow lid labelled “Cells from CSF”   **Dispose of empty vials – Do not ship or re-use them** | 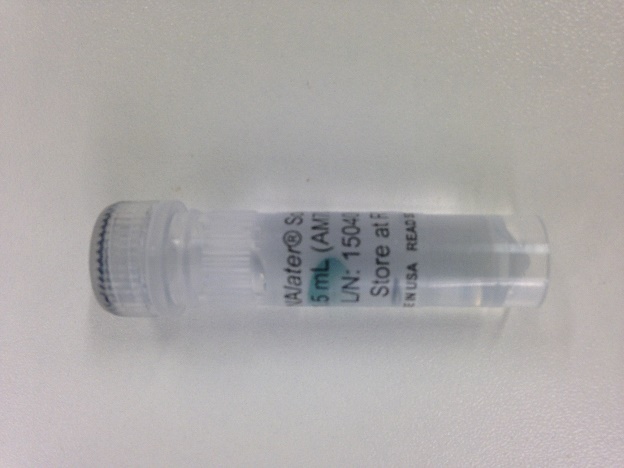  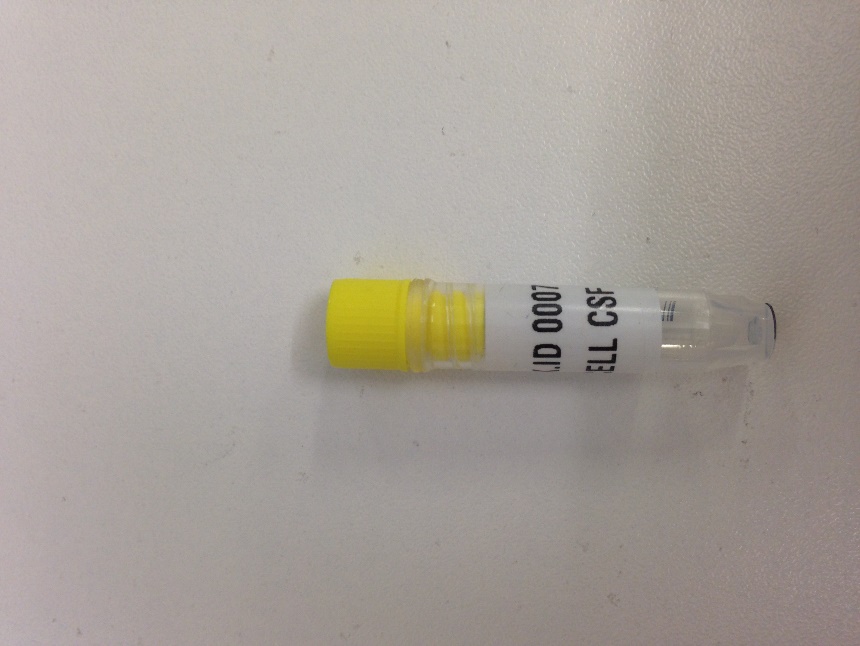 |
| **Sample Storage and Shipment** | 13. Immediately after processing freeze CSF aliquots and the resuspended cells in your -80°C freezer.Ensure samples are stored upright and all lids are secure   Plasma, Serum and CSF do not need to be stored in the freezer at the same time – if waiting for the blood to be ready will cause a delay, then store the CSF in the freezer first, rather than waiting.  If there will be any delay in getting the samples into the freezer then they can be kept in dry ice for a short period of up to 5 minutes. Please document this on the worksheet or source notes to explain how the samples were stored if not transferred immediately to the freezer. | FREEZE AT -80°C AND SHIP AFTER A MINIMUM OF 3 MONTHS, AND WHEN YOU HAVE AT LEAST 5 SAMPLES |
|  | Details of CSF processing are recorded on the **CSF** eCRF, ‘CSF processing’ box (Screen shot 2).  Record the following parameters;  Start time of CSF processing  End time of CSF processing  CSF tube rack ID  CSF aliquot tube ID and number of cryovials  Cells from CSF tube ID  Date and time the samples are stored  Any discrepancies in ID must be explained bearing in mind the ID is the only way to reconcile samples with participants | 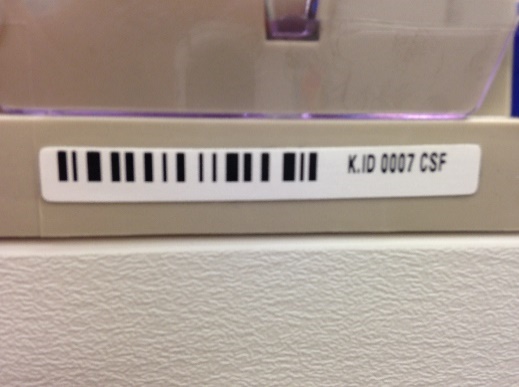  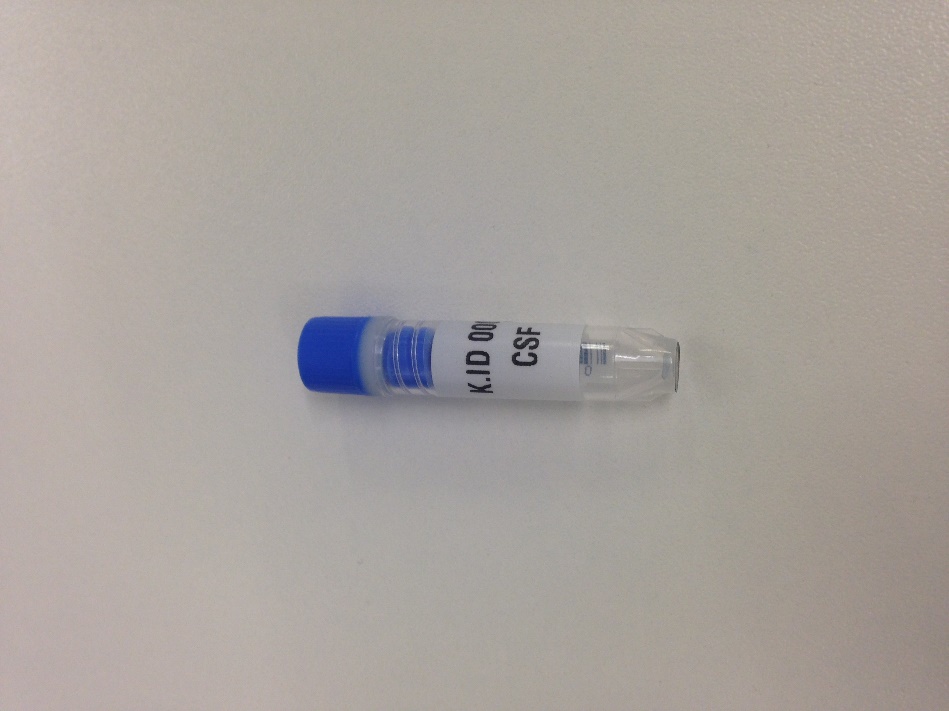  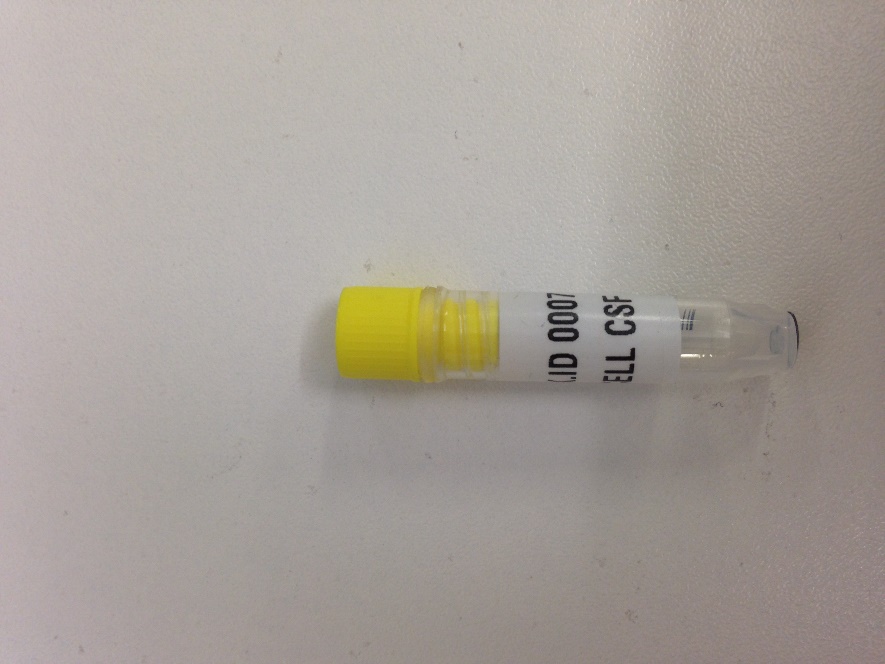 |

Blood processing

| **Sample Collection** | 1. Gently invert each tube 10 times immediately after collection, and place on wet ice |  |
| --- | --- | --- |
|  | 2. Samples transported immediately to laboratory for processing. | 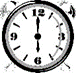  Processing must start within 15 minutes of sample collection |
|  | 3. Lab to receive 4 x 10 ml blood in lithium heparin tubes | 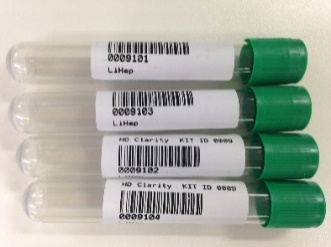 |
| **Sample Processing** | 4. Note the following for later entry into the eCRF, or enter directly:   Lithium heparin tube IDs  Plasma aliquot tube ID  Start time of plasma processing | 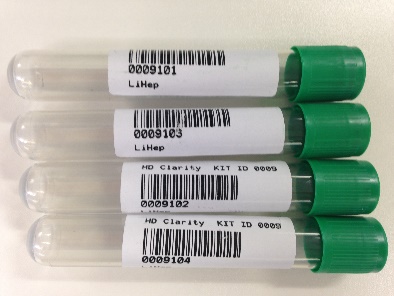  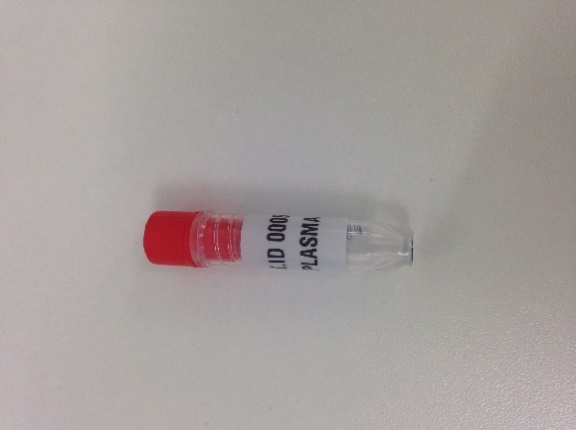 |
|  | 5. Spin lithium heparin tubes at **1300×g for 10 min at 4°C** immediately on arrival |  |
|  | 6. Discard any tubes whose plasma is pink due to haemolysis. In the unlikely event that they are all pink then use all of the tubes but clearly label the sample as contaminated. |  |
|  | 7. Combine the supernatant in one tube labelled “plasma” and mix by inverting 10 times. Place on wet ice. | 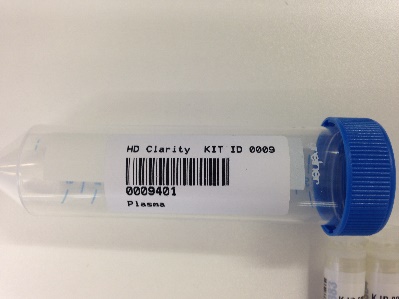 |
|  | 8. Aliquot the plasma into 300 µl cryovials labelled ‘plasma’ using a sterile individually wrapped polypropylene 1ml pipette tip 9. Plasma aliquots must have red lids. Any samples that do not have the expected colour lid will be discarded by BioRep.   **Dispose of empty vials – Do not ship or re-use them** | Dispose of empty vials – do not ship!  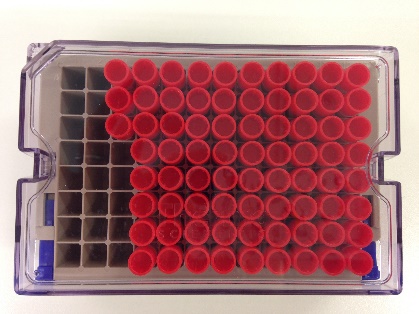 |
| **Sample Storage and Shipment** | 10. Freeze samples on dry ice and store at -80°C   Ensure samples are stored upright and all lids are secure | FREEZE AT -80°C AND SHIP AFTER A MINIMUM OF 3 MONTHS, AND WHEN YOU HAVE AT LEAST 5 SAMPLES |
|  | 11. Record the following on the Blood Processing’ tab in the eCRF (Screen Shot 4);   LiHep tube ID  Processing start time  Plasma aliquot tubes ID  Plasma aliquot tube count  Time plasma processing is completed  Time of frozen storage (if serum and plasma times of freezing are different then it is the time of freezing the **plasma** which is most important to record in the EDC) |  |

#### Supplementary appendix 3 - HDClarity Investigators

**Central Coordination**

University College London: Edward J Wild (Chief Investigator), Gail Owen (Study Manager), Filipe B Rodrigues (Quality Control Officer), Katarzyna Schubert (Study Coordinator), Seema Maru (Study Coordinator), Alexander Lowe (Research Assistant), Stefanie Gosling (former Study Coordinator).

CHDI Foundation: Robi Blumenstein (President), Cristina Sampaio (Chief Clinical Officer), Eileen Neacy (Chief Operating Officer), Swati Sathe (Medical Director, Clinical Research), Anka G Ehrhardt (Director, Bio Fluid Clinical Research), Elena Pak (Clinical Research Program Manager and Study Lead), Shilpa Deshpande (Director, Clinical Operations), Sherry Lifer (Director, Contract Finance & Operations), Julia Keklak (Clinical Program Manager, Biorepository), Dipinder Kaur (former Clinical Biorepository Program Manager), Jamie Levey (Co-Director, Clinical Research Platform), Olivia Handley (Enroll-HD Global Project Manager), Jenny Townhill (Enroll-HD Trial Manager), Mette Gilling (Enroll-HD Scientific Project Manager).

**Study Sites (February 2016 to September 2019)**

*Center Movement Disorders, CA:*

Mark Guttman (Principal Investigator), Bhavpreet Dam (Sub-Investigator), Ragani Srinivasan (Sub-Investigator), Ben Safa (Sub-Investigator), Keith Tanner (Sub-Investigator), Fahad Alam (Sub-Investigator), Jonielyn Carlos (Study Coordinator), Teena Kailasanathan (Study Coordinator), Marijana Pajic (Study Coordinator), Theresa Moore (Study Coordinator), Susan Whyte (Study Coordinator), Tania Mani (Study Coordinator), Marie Villagonzalo (Study Coordinator), Kim Thompson (Study Coordinator).

*University British Columbia, CA:*

Blair Roland Leavitt (Principal Investigator), Lynn Alison Raymond (Sub-Investigator), Mike Adurogbangba (Study Coordinator), Fabricio J Pio (Rater) Emma Peachey (Research Assistant), Jonathan Squires (Sub-Investigator), Valerie O’Neill (Study Coordinator), Tuan Le (Study Coordinator), Rachel Wan (Research Assistant), Devine Calanog (Research Assistant) and Tariq Aziz (Lab Manager).

*George Huntington Institute, DE:*

Ralf Reilmann (Principal Investigator), Stefan Bohlen (Deputy / Sub-Investigator), Anabel Ruesenberg (Sub-Investigator), Anja Kletsch (Study Coordinator/Rater), Laura Spital (Study Coordinator/Rater), Paula Raulet (Study Coordinator/Rater).

*St Josef And Elisabeth Hospital, DE:*

Carsten Saft (Principal Investigator), Sarah Maria von Hein (Sub-Investigator), Jannis Achenbach (Sub-Investigator), Barbara Kaminski (Study Coordinator/Study Nurse), Daniela Kaminski (Study Nurse).

*University Hospital Erlangen, DE:*

Jürgen Winkler (Representative Principal Investigator 07-MAR-2018 until 14-JAN-2019; Principal Investigator since 15-JAN-2019), Zacharias Kohl (Principal Investigator 07-Mar-2018 until 14-JAN-2019), Franz Marxreiter (Sub-Investigator 07-MAR-2018 until 14-JAN-2019; Representative Principal Investigator since 15-JAN-2019), Martin Regensburger (Sub-Investigator), Susanne Seifert (Study Coordinator), Holger Meixner (Laboratory Technical Staff), Jasmin Burczyk (Study Administration), Pia-Marie Pryssok (Study Nurse)

*University Hospital Ulm, DE:*

*J*an Lewerenz (Principal Investigator)

*BirmSolNHSFounTrust, UK:*

Hugh Rickards (Principal Investigator), Diana Crossley (Sub-Principal Investigator), Aaron Sturrock (Sub-Principal Investigator), Jennifer De Souza (Study Coordinator/Rater), Theresa Brady (Research Nurse), Anna Finnegan (Research Nurse), Samantha Timmis (Research Nurse), Maria Bandeira (Data Officer), Tracy Soulsby (Data Officer), Nula Kelly (Research Nurse), Melissa Wardale (Lab Manager), Fahd Niaz (Data Officer).

*GreatGlasgowHealthBoard, UK:*

Stuart Ritchie (Principal Investigator), Stuart Affleck (Sub-Investigator), Paul Gallagher (Sub-Investigator), Stephanie Cowan (Sub-Investigator), Sarah Martin (Sub-Investigator), Shoshana Cross (Sub-Investigator), Gillian Scott (Sub-Investigator), Craig Patrick (Sub-Investigator), Catherine Deith (Study Coordinator), Carol Malcolmson (Study Coordinator), Murray Sutherland (Research Nurse), Scott Farmer (Research Nurse), Lanah Dunsmuir (Research Nurse), Anne Lewis (Lab Manager).

*LeedsTeachHospTrust, UK:*

Jeremy Cosgrove (Principal Investigator), Callum Schofield (Study Coordinator), Alan Liu (Research Nurse), Helena Baker (Biomedical Scientist), Jodie Sedgwick (Biomedical Scientist).

StGeorgeHealthTrust, *UK*:

Nayana Lahiri (Principal Investigator), Bhavini Patel (Sub-Investigator), Sally Goff (Study Coordinator), Uruj Anjum (Study Coordinator), Chandni Patel (Study Coordinator).

University Cambridge, *UK*:

Roger Barker (Principal Investigator), Thomas Stoker (Sub-Investigator), Katie Andresen (Study Coordinator/Rater).

*University College London, UK:*

Edward J Wild (Chief Investigator/Principal Investigator), Filipe Brogueira Rodrigues (Sub-Investigator), Lauren M Byrne (Study Coordinator/Rater), Rosanna Tortelli (Sub-Investigator), Peter McColgan (Sub-Investigator), Mike Flower (Sub-Investigator), Carlos Estevez-Fraga (Sub-Investigator), Paul Zeun (Sub-Investigator), Carolin Koriath (Sub-Investigator), Edwina Saunders (Research Nurse), Mila Resuello-Dauti (Research Nurse), Laura Hennelly (Research Nurse), Nuria Mora Morell (Research Nurse), Mark Elliot (Nurse Assistant), Rhoda Castaneda (Study Coordinator), Martha S. Foiani (Research Technician), Jamie Toombs (Research Technician), Elena Veleva (Research Technician), Michael Chou (Research Technician).

*JohnsHopkinsUniv, US:*

Jee Bang (Principal Investigator), Christopher Ross (Sub-Investigator), Kia E Ultz (Study Coordinator/Rater), Jacqueline V Bran (Study Coordinator), Eka Chighladze (Lab Technician), Priyanka Rauniyar (Lab Technician), Chelsy Eddings (Lab Technician).

*WakeForestUniv, US:*

Francis Walker (Principal Investigator), Clarisse Goas (Sub-Investigator), Victoria Hunt (Research Nurse), Christine O’Neill (Study Coordinator/Rater), Jessica Bargoil (Study Coordinator/Rater), Sara Byerly (CRU Manager), Cathy Gilkey (Lab Technician), LuAnn Mascorro (Lab Technician).

*UnivTexasHlthCntrHous, US:*

Erin Furr-Stimming (Principal Investigator), David Hunter (Sub-Investigator), Beth Latham (Study Coordinator/Rater), Jamie Sims (Study Coordinator/Rater), Brittany Duncan (Study Coordinator/Rater).
